# A panel of synapse assembly genes as a biomarker for Gliomas

**DOI:** 10.1101/19011114

**Authors:** Xiangwen Ji, Hongwei Zhang, Qinghua Cui

**Affiliations:** Department of Biomedical Informatics, Department of Physiology and Pathophysiology, Center for Noncoding RNA Medicine, MOE Key Lab of Cardiovascular Sciences, School of Basic Medical Sciences, Peking University, 38 Xueyuan Rd, Beijing, 100191, China; Department of Neurosurgery, Sanbo Brain Hospital, Capital Medical University, Beijing, 100093, China

## Abstract

Gliomas are the most common primary brain cancers. In recent years, IDH mutation and 1p/19q codeletion have been suggested as biomarkers for the diagnosis, treatment and prognosis of gliomas. However, these biomarkers are only effective for a part of glioma patients and thus more biomarkers are still emergently needed. Recently, an electrochemical communication between normal neurons and glioma cells by neuro-glioma synapse has been reported. Moreover, it was discovered that breast-to-brain metastasis tumor cells have pseudo synapses with neurons and these synapses were indicated to promote tumor progression and metastasis. Based on the above observations, we first curated a panel of 66 SA genes and then proposed a metric, SA score, to quantify the synapseness for each sample of 12 glioma gene expression datasets from TCGA, CGGA, and GEO. Strikingly, SA score showed excellent predictive ability for the prognosis, diagnosis, and grading of gliomas. Moreover, being compared with the two established biomarkers, IDH mutation and 1p/19q codeletion, SA score was demonstrated independent and better predictive performance. In conclusion, this study revealed that SA genes contribute to glioma formation and development, and proposed a quantitative method, SA score, as an efficient biomarker for monitoring gliomas.

## Introduction

Brain and other nervous system cancers are estimated to take up 1.4% of new cancers but 2.9% of cancer deaths in 2019^1^. Gliomas are the most frequent cancers of these cancers, including astrocytoma (including glioblastoma), oligodendroglioma, ependymoma, oligoastrocytoma (mixed glioma), malignant glioma, not otherwise specified (NOS), and a few rare histologies^2^. World Health Organization (WHO) classified gliomas into grades I to IV, and introduced biomarkers of IDH mutation and 1p/19q codeletion in 2016 edition^3,4^. Glioblastoma (WHO grade IV) accounts for about half of gliomas, with a median survival of less than two years^2,5^. Gliomas with lower grade have a diverse prognosis, either progressing to be as poor as glioblastoma or living more than 10 years after effective treatment^6^.

Over the years, with the fast improvement of omics and big data technology, RNA sequencing has been developing towards lower cost and higher throughput, producing a large amount of biological and medical data, which provides great convenience for life science research^7,8^. Impelled by advantage of big data analysis, numerous biomarkers have been found in the diagnosis and prognosis of gliomas^9,10^. In addition, single-cell sequencing showed more detailed landscapes of gliomas^11-13^. Gene set enrichment analysis (GSEA) provides a facility to extract effective information from a large number of RNA expression data^14^. Moreover, single sample GSEA (ssGSEA) can calculate without group information and give every sample an enrichment score ^15^. The Biomarkers such as IDH mutation and 1p/19q codeletion provided helps for monitoring the development and prognosis of gliomas but are only effective for a part of patients^16^. Therefore, given the enormous severity of gliomas, more biomarkers are emergently needed.

It is recently reported that neuron and glioma have electrochemical communication through AMPA receptor-dependent synapses between presynaptic neurons and postsynaptic glioma cells^17,18^. These observations suggest that the neural synaptic electrochemical connections promote glioma progression. Simultaneously, an appearance of glutamatergic ‘pseudo-tripartite’ synapses between breast-to-brain metastasis tumor cells and neurons was observed^19^. Based on these anatomical and cytological findings, we hypothesized that the synapse assembly (SA) genes can be used as a biomarker for glioma prognosis. To confirm this hypothesis, here we first curated a list of genes involved in SA and then performed ssGSEA analysis for glioma gene expression datasets from the Cancer Genome Atlas (TCGA), the Chinese Glioma Genome Atlas (CGGA), and the Gene Expression Omnibus (GEO). Interestingly, the tumor synapses may be involved in immune escape and multiple molecular pathways. Strikingly, SA was found to be an independent and effective biomarker for gliomas.

## Results

### The panel of synapse assembly genes serves as a novel biomarker for gliomas

In order to investigate whether SA can be biomarker for glioma patients, we first curated a list of SA genes. As a result, 66 genes were collected (**Supplementary File 1**). Then we performed ssGSEA to TCGA and CGGA datasets for survival analysis. The results showed glioma patients with higher SA scores have longer overall survival time (**Fig 1**). Cox regression analysis also shows the same results (**Table 1**). Moreover, patients with higher WHO grade have significantly lower SA scores (**Fig. 2**), which agrees with the survival analysis. In addition, it is deserved to be mentioned that SA score shows an ability to distinguish between glioma and normal brain tissue, which reveals a potential diagnostic application of SA (**Fig. 2e-g, Supplementary Fig. 1**; AUCs of ROC analysis for GSE4290, GSE16011, and GSE50161 datasets are 0.91, 0.99, and 0.93, respectively).

**Table 1.**
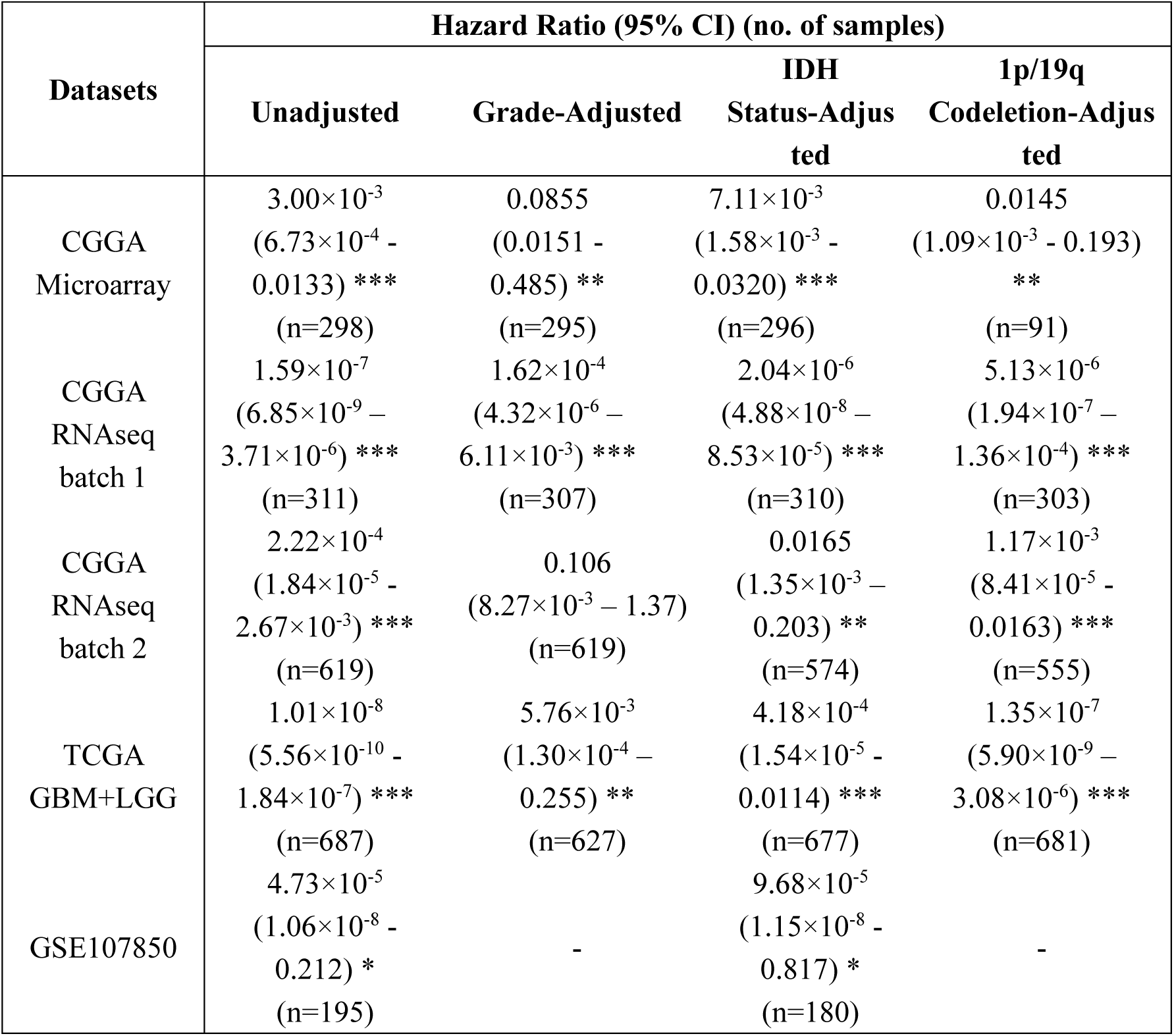
The predictive ability of SA score adjusted using WHO grade, IDH mutation and 1p/19q codeletion. Hazard rate (HR) and 95% confidence interval (95% CI) of synapse assembly (SA) score using univariate and multivariate Cox proportional hazards regression models for gliomas were shown. HR with 95% CI that does not include one is considered significant. * p<0.05, ** p<0.01, *** p<0.001.

| Datasets | Hazard Ratio (95% CI) (no. of samples) |  |  |  |
| --- | --- | --- | --- | --- |
|  | Unadjusted | Grade-Adjusted | IDH<br>Status-Adjusted | 1p/19q<br>Codeletion-Adjusted |
| CGGA<br>Microarray | $3.00 \times 10^{-3}$<br>($6.73 \times 10^{-4}$ -<br>0.0133) ***<br>(n=298) | 0.0855<br>(0.0151 -<br>0.485) **<br>(n=295) | $7.11 \times 10^{-3}$<br>($1.58 \times 10^{-3}$ -<br>0.0320) ***<br>(n=296) | 0.0145<br>($1.09 \times 10^{-3}$ - 0.193)<br>**<br>(n=91) |
| CGGA<br>RNAseq<br>batch 1 | $1.59 \times 10^{-7}$<br>($6.85 \times 10^{-9}$ -<br>$3.71 \times 10^{-6}$) ***<br>(n=311) | $1.62 \times 10^{-4}$<br>($4.32 \times 10^{-6}$ -<br>$6.11 \times 10^{-3}$) ***<br>(n=307) | $2.04 \times 10^{-6}$<br>($4.88 \times 10^{-8}$ -<br>$8.53 \times 10^{-5}$) ***<br>(n=310) | $5.13 \times 10^{-6}$<br>($1.94 \times 10^{-7}$ -<br>$1.36 \times 10^{-4}$) ***<br>(n=303) |
| CGGA<br>RNAseq<br>batch 2 | $2.22 \times 10^{-4}$<br>($1.84 \times 10^{-5}$ -<br>$2.67 \times 10^{-3}$) ***<br>(n=619) | 0.106<br>($8.27 \times 10^{-3}$ - 1.37)<br>(n=619) | 0.0165<br>($1.35 \times 10^{-3}$ -<br>0.203) **<br>(n=574) | $1.17 \times 10^{-3}$<br>($8.41 \times 10^{-5}$ -<br>0.0163) ***<br>(n=555) |
| TCGA<br>GBM+LGG | $1.01 \times 10^{-8}$<br>($5.56 \times 10^{-10}$ -<br>$1.84 \times 10^{-7}$) ***<br>(n=687) | $5.76 \times 10^{-3}$<br>($1.30 \times 10^{-4}$ -<br>0.255) **<br>(n=627) | $4.18 \times 10^{-4}$<br>($1.54 \times 10^{-5}$ -<br>0.0114) ***<br>(n=677) | $1.35 \times 10^{-7}$<br>($5.90 \times 10^{-9}$ -<br>$3.08 \times 10^{-6}$) ***<br>(n=681) |
| GSE107850 | $4.73 \times 10^{-5}$<br>($1.06 \times 10^{-8}$ -<br>0.212) *<br>(n=195) | - | $9.68 \times 10^{-5}$<br>($1.15 \times 10^{-8}$ -<br>0.817) *<br>(n=180) | - |

**Fig. 1.**
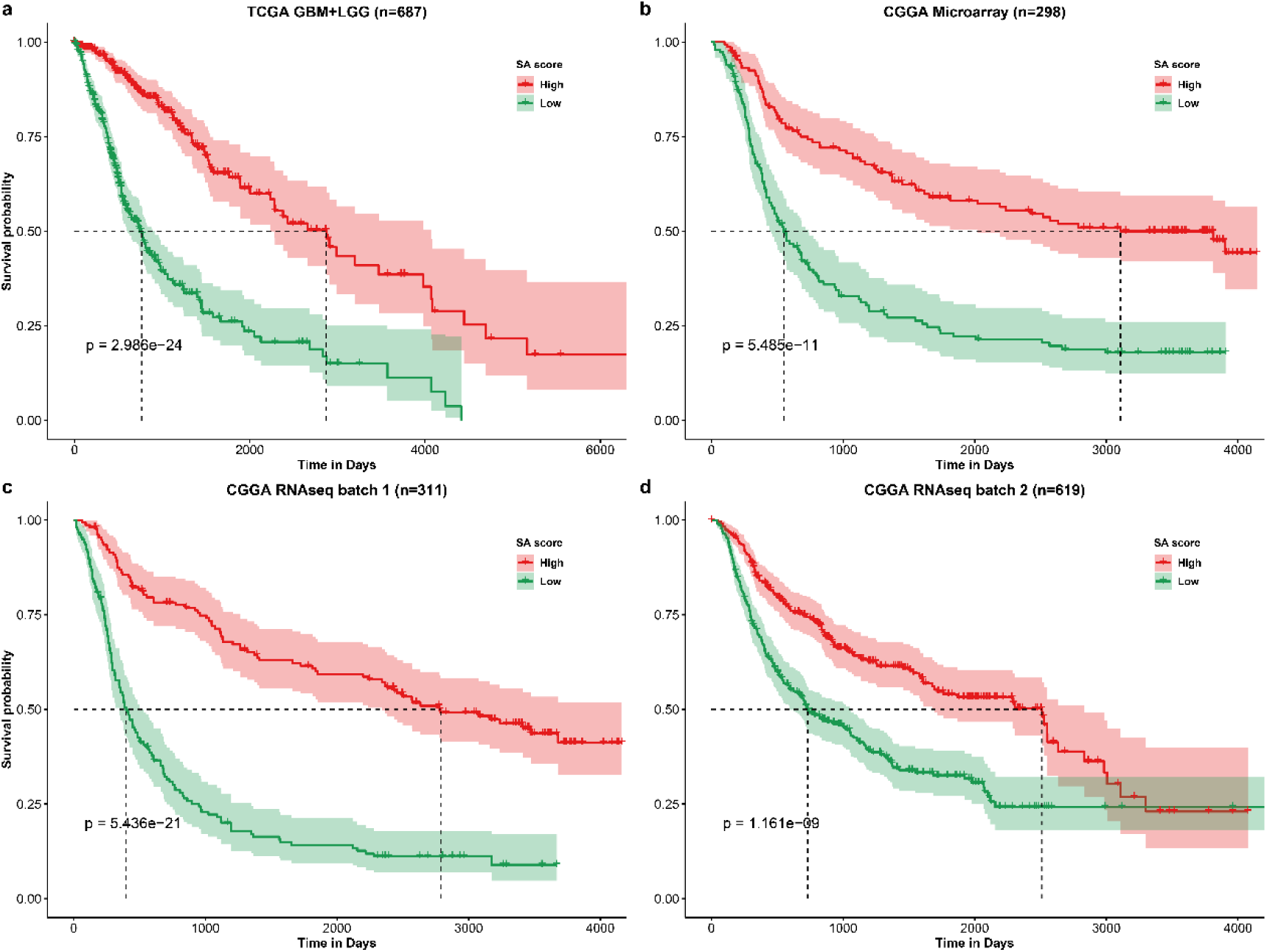
Kaplan-Meier curve of overall survival. (a) TCGA lower grade glioma (LGG) and glioblastoma multiforme (GBM). (b) CGGA Microarray. (c) CGGA RNAseq batch 1. (d) CGGA RNAseq batch 2. Group was separated by the median value of SA scores. Differences between two curves were estimated by log-rank test.

**Fig. 2.**
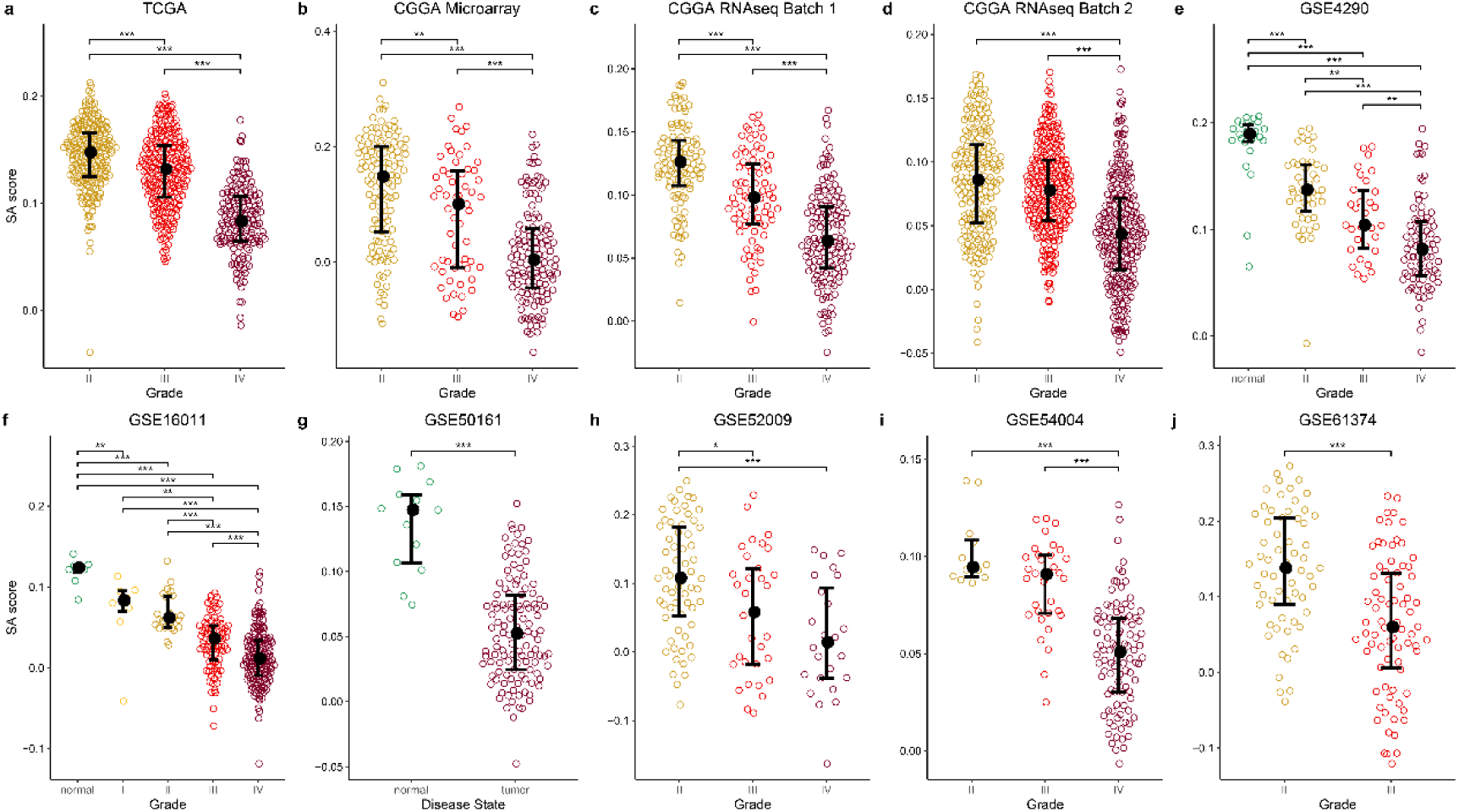
SA scores were significantly lower in higher grade gliomas. (a) TCGA lower grade glioma (LGG) and glioblastoma multiforme (GBM). (b) CGGA Microarray. (c) CGGA RNAseq batch 1. (d) CGGA RNAseq batch 2. (e) GSE4290. (f) GSE16011. (g) GSE50161. (h) GSE52009. (i) GSE54004. (j) GSE61374. Significances of difference between two groups were analyzed by two-side Wilcoxon rank sum test. * p<0.05, ** p<0.01, *** p<0.001.

### Comparison of SA score with established biomarkers

IDH mutation and 1p/19q codeletion are two established biomarkers for gliomas. Both biomarkers provided great helps for monitoring glioma development but both are effective on only a part of patients. Therefore, it is interesting to explore whether SA score is an independent biomarker and whether SA score is better than the established biomarkers or not. For doing so, we first analyzed the relationship of SA scores with IDH mutation and 1p/19q codeletion status. We found that IDH-mut gliomas were associated with significantly higher SA scores than IDH-wt ones (**Fig. 3a-d**). And 1p/19q codeletion gliomas represent a higher SA scores than non-codeletion ones (**Fig. 3e-h**). Moreover, after moving the effects of the two established biomarkers using multivariate Cox regression model, we revealed that SA score is an independent biomarker for predicting prolonged overall survival in gliomas (**Table 1**). In addition, the grading ability of SA score is also independent to IDH mutation and 1p/19q codeletion (**Supplementary Fig. 2**). Finally, we compared the predictive performance of SA score, IDH mutation and 1p/19q codeletion status (**Table 2**). In most instances, SA score outperforms IDH mutation and 1p/19q codeletion.

**Table 2.** The predictive ability for survival prognosis of SA score compared with IDH mutation and 1p/19q codeletion status. Patients were divided into two groups according to the biomarker, and the difference (p value of log-rank test) between two K-M curves was calculated. The threshold of SA score is determined by the best separation it can perform. The better results are highlighted in bold.

| Datasets | SA score | IDH Status | SA score | 1p/19q Status |
| --- | --- | --- | --- | --- |
| CGGA<br>Microarray | **2.387**$\times 10^{-13}$ | 6.159$\times 10^{-10}$ | **1.341**$\times 10^{-5}$ | 4.176$\times 10^{-5}$ |
| CGGA RNAseq<br>batch 1 | **6.654**$\times 10^{-25}$ | 2.859$\times 10^{-12}$ | **2.761**$\times 10^{-24}$ | 4.156$\times 10^{-15}$ |
| CGGA RNAseq<br>batch 2 | 1.012$\times 10^{-11}$ | **6.639**$\times 10^{-29}$ | 6.103$\times 10^{-10}$ | **9.095**$\times 10^{-14}$ |
| TCGA<br>GBM+LGG | 3.745$\times 10^{-29}$ | **1.363**$\times 10^{-77}$ | **4.786**$\times 10^{-30}$ | 1.194$\times 10^{-13}$ |
| GSE107850 | **1.241**$\times 10^{-3}$ | 1.633$\times 10^{-2}$ | - | - |

**Fig. 3.**
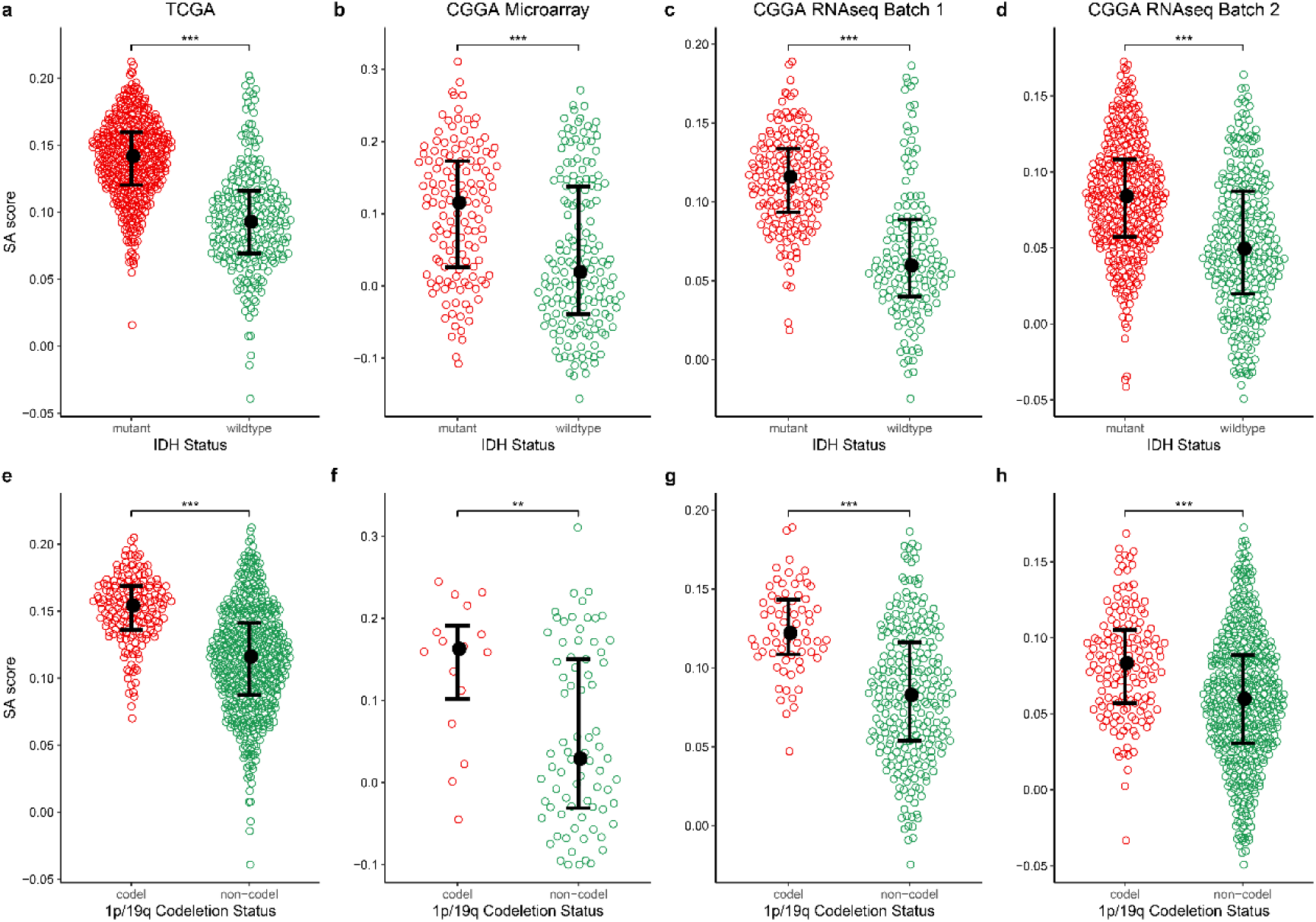
SA scores were associated with IDH mutation and 1p/19q codeletion status. (a, e) TCGA lower grade glioma (LGG) and glioblastoma multiforme (GBM). (b, f) CGGA Microarray. (c, g) CGGA RNAseq batch 1. (d, h) CGGA RNAseq batch 2. Significances of difference were analyzed by two-side Wilcoxon rank sum test. ** p<0.01, *** p<0.001.

## Discussion

Given the recently revealed roles of neuro-glioma synapse in glioma development, here we curated a panel of 66 synapse assembly (SA) genes and proposed the SA score as a biomarker for the prognosis, grading, and diagnosis of gliomas. The SA score was validated by over 3000 samples of 12 datasets from TCGA, CGGA, and GEO.

To confirm whether the SA score does represent the process of synapse assembly or not, we performed Spearman’s correlation analysis between SA score and expressions of synapse receptor genes ^17^ (**Supplementary Fig. 3a**). The result showed that a number of genes, especially AMPA (α-amino-3-hydroxy-5-methyl-4-isoxazole propionic acid) receptor genes (not contain in SA genes), which were focused in recent studies^17,18^, have a positive correlation with SA scores. Then, we applied pathway analysis to genes with significant correlation with SA scores (<u>Methods</u>). As a result, the positively correlated genes were significantly enriched in synaptic related pathways (**Supplementary Fig. 3b, c**) and cell adhesion molecules (CAMs, hsa04514) in neural system (**Supplementary Fig. 4**), suggesting that the calculated SA score can represent the process of synapse assembly.

In spite of its ability as glioma biomarker for the identified SA gene panel, it should be especially noted that the result seems opposite to existed knowledge. That is, it was reported that neuro-glioma synapse could promote tumor progression and metastasis^17-19^, which thus can infer that SA should result in a poorer prognosis, however, we revealed it is associated with a better but not poorer prognosis. Molecular processes may play different roles in various cells, organs and diseases. For example, as an important discovery in glioma research, IDH mutation is identified as one of the early events of glioma, and the epigenetic changes caused by IDH mutation are considered as a main tumor driver ^20^. Nevertheless, clinical studies have found that IDH mutation can lead to a longer survival time ^21^. Similarly, immunotherapy, which has been widely used, was criticized for producing serious side effects ^22,23^. These instances suggest that the SA gene panel could also have multiple aspects.

To better understanding the molecular processes of the SA genes, we sought to pathway analysis. Besides the proliferative effect suggested in previous studies, we revealed that neuro-glioma synapses may also show a participation in the immune response, cyclic adenosine monophosphate (cAMP, hsa04024) and nuclear factor kappa-B (NF-κB, hsa04064) pathway (**Supplementary Fig. 3b, c**). The SA genes are related to the activation of cAMP pathway (**Supplementary Fig. 3c**, 5), which has been proved to be involved in the synaptic plasticity process, including AMPA receptor trafficking, which may be a reason why glioma cells form synapses and produce AMPA receptors^24^. Moreover, cAMP can also activate phosphoinositide 3-kinases (PI3K)/protein kinase B (PKB, also known as Akt), thus promoting cell growth, survival and proliferation, which may be one of the explanations for the proliferation promotion effect of the newly discovered neuro-glioma synapse ^25^. Since the synaptic structure was also observed in the breast-to-brain cancer cells, we hypothesized that it was not the cell origin but the environment that determined this morphological change in tumor. It is worth mentioning that the cAMP pathway can be activated by neurotransmitters and Ca^2+^, which is extensively distributed in synaptic cleft and easily accessible for glioma^26-28^.

The NF-κB signaling pathway and CAMs of immune system were also found to be vastly enriched with genes negatively associated with SA (**Supplementary Fig 3c, 4, 6**). Previous studies on NF-κB are mainly focused on immune cells, which are involved in cell activation and survival, as well as the production of cytokines^29,30^. In tumors, the stimulation of NF-κB signal leads to up-regulation of anti-apoptotic genes that allow tumors to tolerate inflammatory environment. Meanwhile, NF-κB induces cytokines and CAMs, which give rise to the recruitment of immune cells to the tumor site^31^. As can be seen, the relationship between NF-κB-caused inflammation and cancer is complex, which can either eliminate cancer cells or lead to immune escape. Additionally, stimulated cAMP could suppress NF-κB activation ^32^. This finding, together with the relationship between NF-κB and CAMs, is in good agreement with our discoveries. We also demonstrated that SA is associated with IDH mutation status, which is a momentous survival risk factor. In a previous study, significantly higher tumor infiltrating lymphocytes (TILs) infiltration and programmed death ligand 1 (PD-L1) expression, which are targets of immune checkpoint inhibitor, were found to be associated with IDH-wt^33^. However, the others have found that IDH-mut tumors show reductions of various immune cells, including microglia, macrophages and polymorphonuclear leukocytes, which are consistent with the decrease of chemokines in tumors^34^.

As it was shown in recent studies^17-19^, the neuro-glioma synapses are involved in cancer proliferation and metastasis, which are beneficial to tumor progression. But reversely, we found that higher SA signifies lower glioma grade and longer overall survival. Analogously, IDH-mut and 1p/19q codeletion are typically biomarkers that promote glioma progression but benefit prognosis. Existing studies have focused on mechanisms that promote glioma, but the reasons for better prognosis are generally reported by clinical studies, such as better chemoradiotherapy sensitivity^35^. We conjectured that SA, IDH mutation and 1p/19q codeletion shared a part of the mechanism that resulted in the observed phenomenon. Apoptosis caused by suppression of NF-κB signaling pathway may also be a motivation. The causations in SA, mutation, pathway and immune response are remained to be explored.

In summary, we found that SA is associated with activation of the cAMP signaling pathway and down-regulation of NF-κB pathway and immune-related CAMs. Although the mechanism is unclear, we revealed that SA genes are involved in glioma process and the proposed SA score is an independent and potentially better biomarker for glioma overall survival, and shows a predictive capacity in different grade gliomas and normal brain tissue, which could be useful in the prognosis, grading and diagnosis of gliomas.

## Methods

### Gene expression datasets and analysis

We obtained sample and clinical information from TCGA (https://portal.gdc.cancer.gov/). RNAseq data of TCGA were downloaded from FireBrowse (http://firebrowse.org/), which were processed and normalized uniformly. Histology type, WHO grade, IDH mutation status and 1p/19q codeletion status were obtained from the study by Ceccarelli et al^36^. CGGA (http://www.cgga.org.cn/) provides tumor gene expression data for thousands of glioma patients (including one microarray and two RNAseq batches), as well as corresponding clinical data. The calculation and presentation of the results will be conducted separately due to different platforms and batches. In addition, glioma microarray gene expression profiling data (GSE4290, GSE16011, GSE50161, GSE52009, GSE54004, GSE61374, GSE107850) were available at GEO (https://www.ncbi.nlm.nih.gov/gds/). Gene expression data were structured with gene symbols as row names, sample ids as column names, duplicate gene symbols were averaged using their median value.

### Single sample gene set enrichment analysis

The human synapse assembly gene set was extracted from the Gene Ontology (GO) by term ‘positive regulation of synapse assembly’ (GO:0051965). The ssGSEA scores were calculated by python (v3.6.8) package gseapy (v0.9.13), which is a python wrapper for GSEA and ssGSEA.

### GO and KEGG pathway analysis

We performed Spearman’s correlation analysis between SA ssGSEA score and gene expression in the TCGA low grade glioma (LGG) and CGGA (3 subdatasets) gliomas datasets. Top 5000 (ordered by rho) significantly positively/negatively correlated genes were selected respectively and the results of four datasets were intersected. R package clusterProfiler^37^ (v3.10.1) was used for GO and Kyoto Encyclopedia of Genes and Genomes (KEGG) pathway enrichment analysis. P values were adjusted by Benjamini and Hochberg methods and background genes were set as genes which exist in all four datasets. KEGG pathway graphs were rendered by R package pathview^38^ (v1.22.3).

### Statistical analysis

Kaplan-Meier (K-M) curves and Cox proportional hazards regression were performed by R package survival (v2.44-1.1) and survminer (v0.4.6). Log rank test was used to calculate the difference between two K-M curves. Significance of difference between two groups of continuous variables was analyzed by two-side Wilcoxon rank sum test. Receiver operating characteristic (ROC) curve and area under ROC curve (AUROC) were processed by R package pROC^39^ (v1.15.3). All statistical significances above were calculated by R (v3.5.2). Spearman’s correlation analysis was applied to evaluate the correlation using python package scipy (v1.2.1). P values < 0.05 were considered significant.

## Data Availability

The datasets supporting the conclusions of this article are available in TCGA repository, https://portal.gdc.cancer.gov/, the FireBrowse repository, http://firebrowse.org/, the CGGA repository, http://www.cgga.org.cn/ and GEO repository, https://www.ncbi.nlm.nih.gov/gds/.
WHO grade, IDH mutation status and 1p/19q codeletion status of TCGA are available in the supplementary file of the published article by Ceccarelli et al, https://doi.org/10.1016/j.cell.2015.12.028.
The 66 SA genes are available in the supplementary file 1.

https://portal.gdc.cancer.gov/

http://firebrowse.org/

http://www.cgga.org.cn/

https://www.ncbi.nlm.nih.gov/gds/

https://doi.org/10.1016/j.cell.2015.12.028

## Author contributions

QC conceived the project. XJ performed the analysis and conducted the experiments. XJ, HZ and QC wrote the manuscript. All authors read and approved the final manuscript.

## Supplementary information

**Supplementary Fig. 1.**
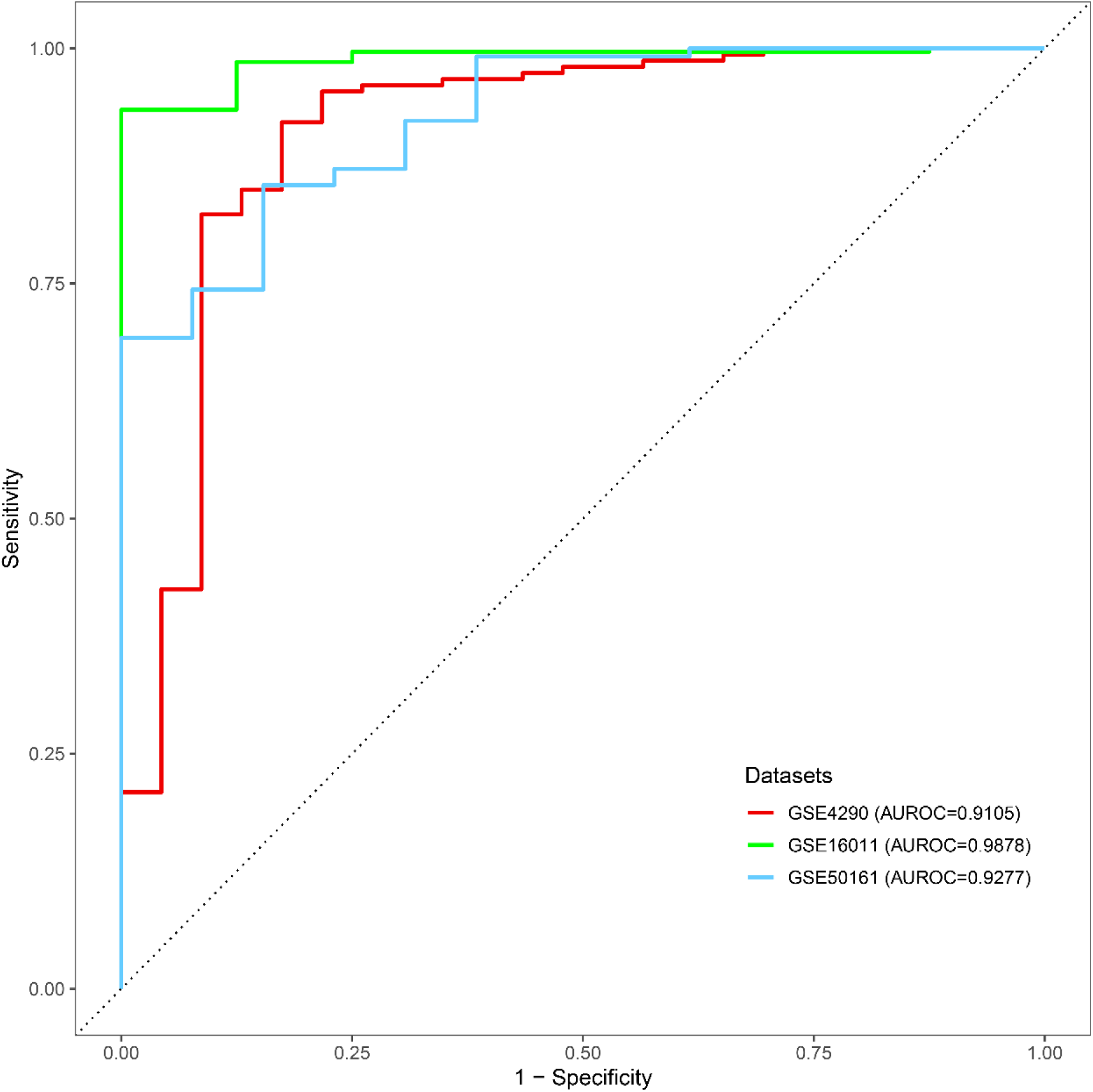
SA scores can discriminate gliomas from normal brain tissue. ROC curves of GSE4290 (red), GSE16011 (green), GSE50161 (blue) from GEO datasets.

**Supplementary Fig. 2.**
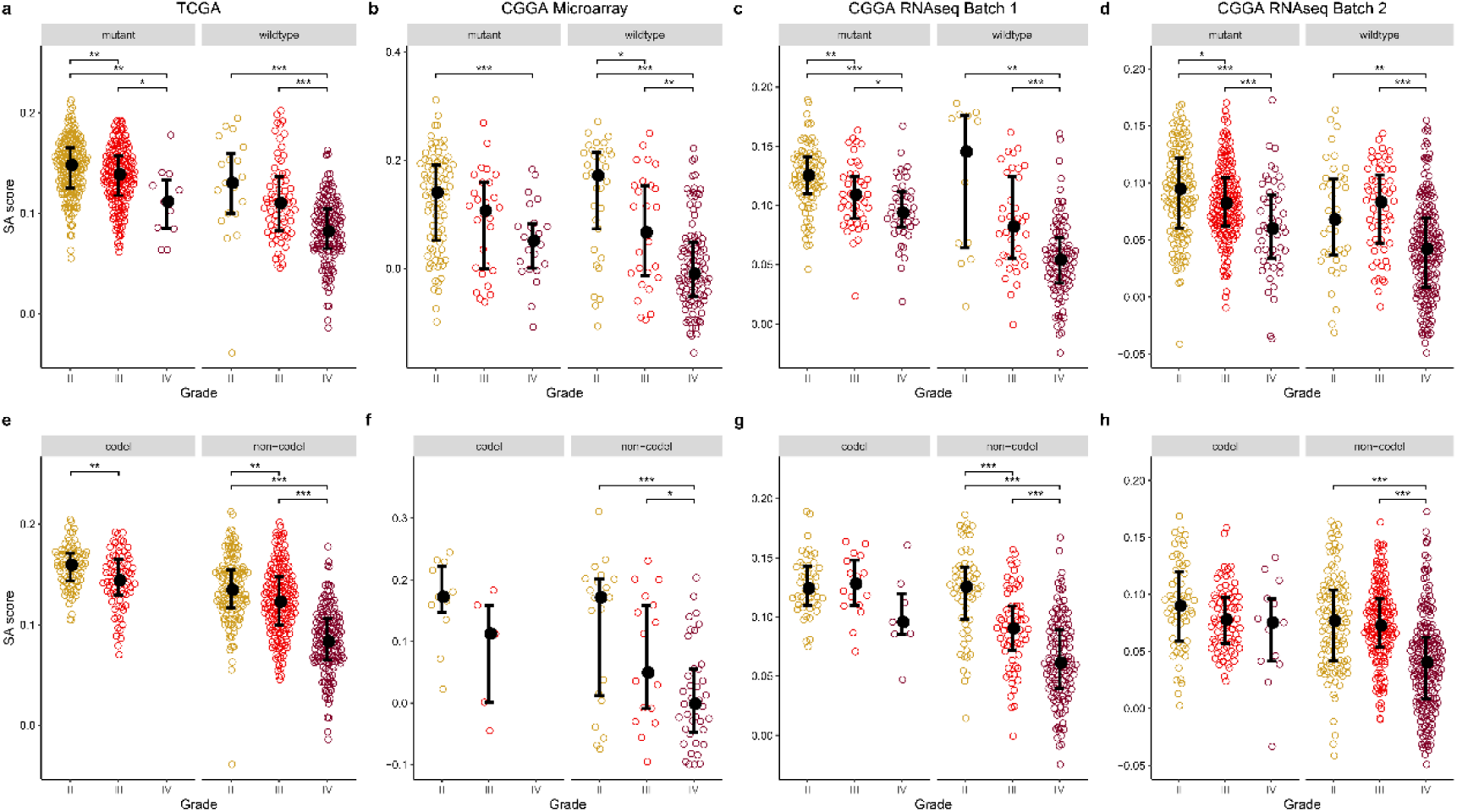
The grading ability of SA score adjusted using IDH mutation and 1p/19q codeletion. (a, e) TCGA lower grade glioma (LGG) and glioblastoma multiforme (GBM). (b, f) CGGA Microarray. (c, g) CGGA RNAseq batch 1. (d, h) CGGA RNAseq batch 2. Significances of difference were analyzed by two-side Wilcoxon rank sum test. * p<0.05, ** p<0.01, *** p<0.001.

**Supplementary Fig. 3.**
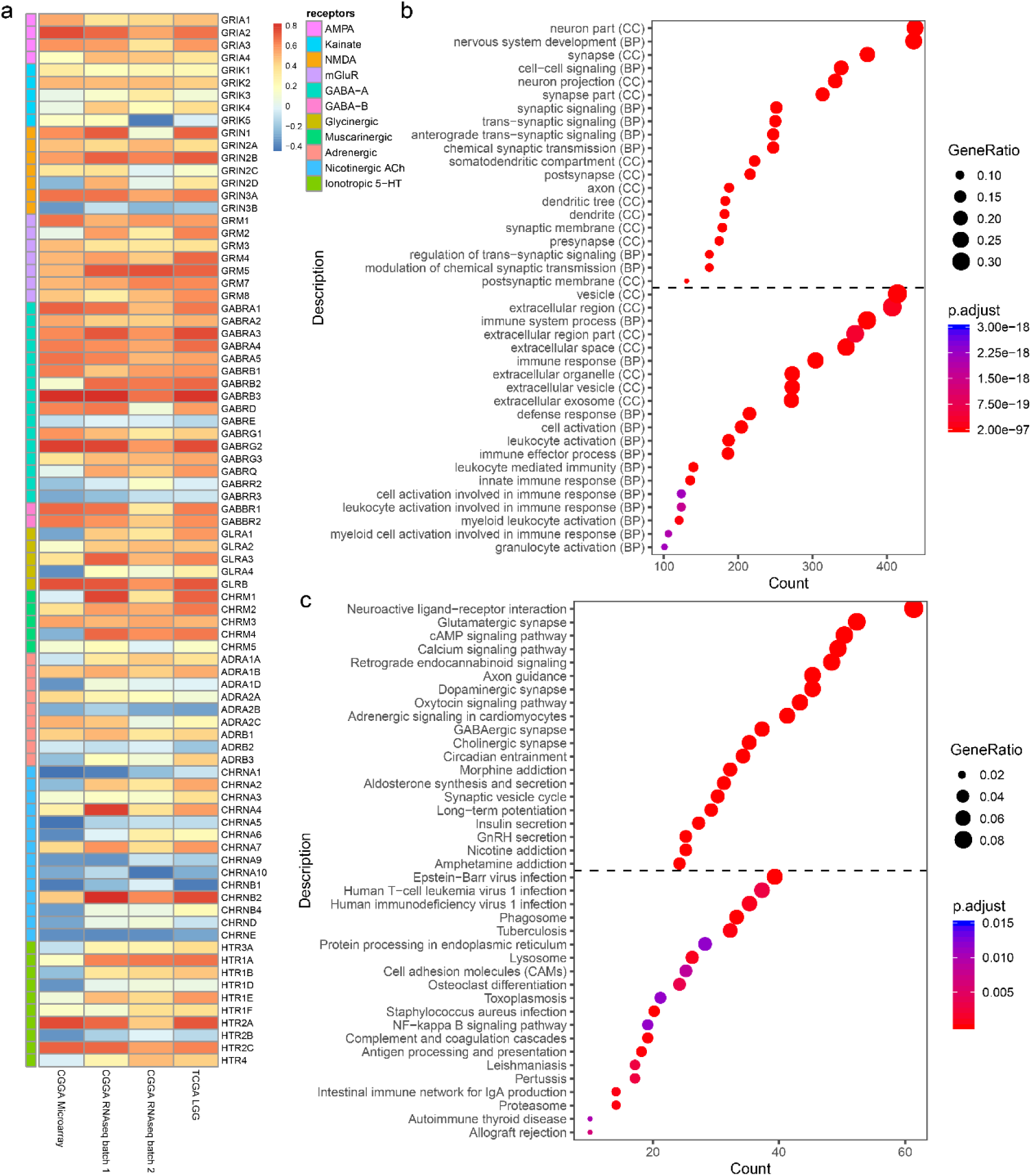
SA score is related with synapse receptors genes and several GO/KEGG pathways. (a) Heatmap shows the Spearman correlation between SA score and synapse receptors genes. (b-c) GO/KEGG enrichment analysis of SA score associated genes. Above the dotted line: enrichment analysis results of positively related genes. Below the dotted line: results of negatively correlated genes. Top 20 results with the lowest p value were shown respectively.

**Supplementary Fig. 4.**
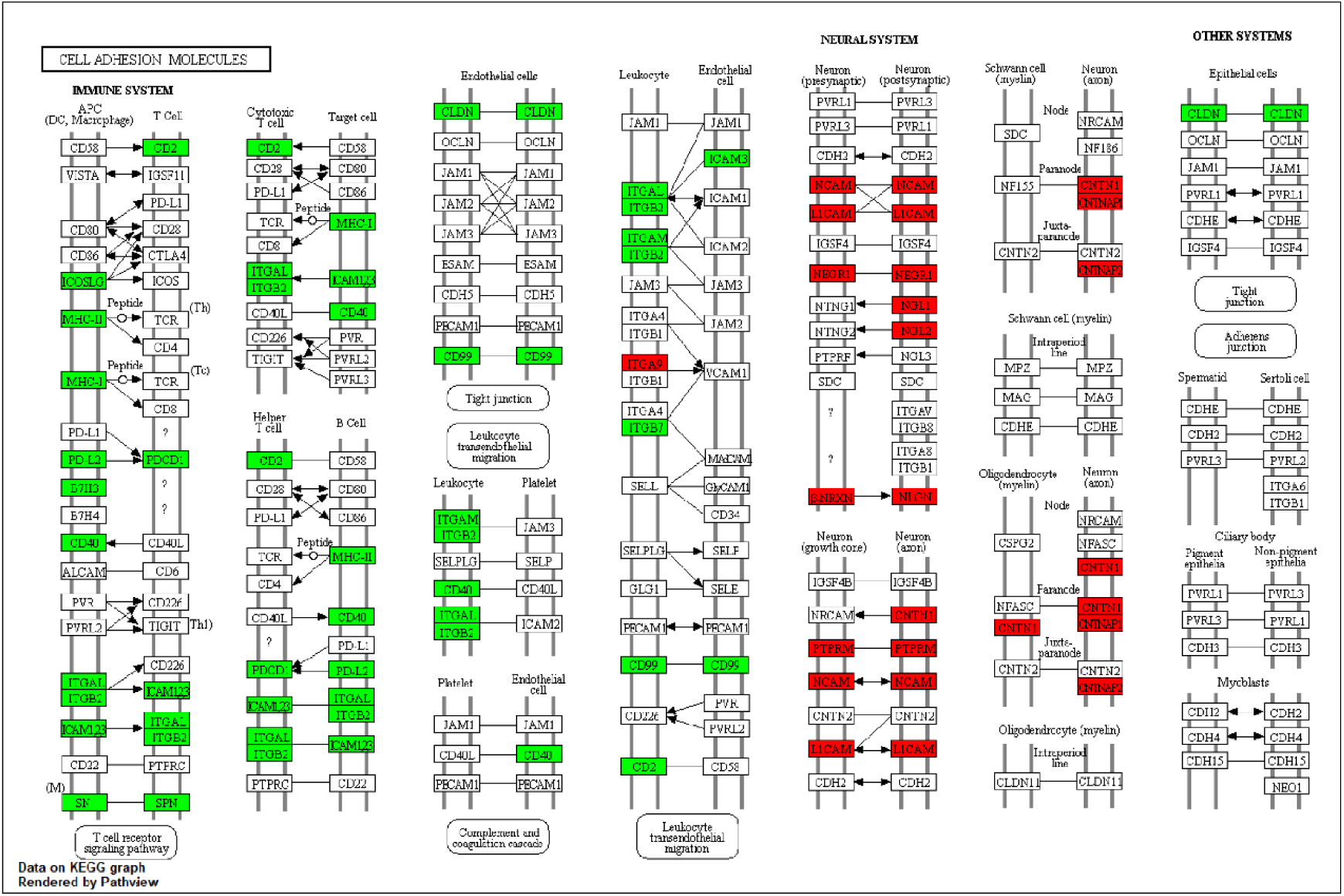
CAMs genes were correlated with SA score. Graph was downloaded from KEGG (hsa04514) and rendered by R package pathview. Red: positive correlation genes. Green: negative correlation genes.

**Supplementary Fig. 5.**
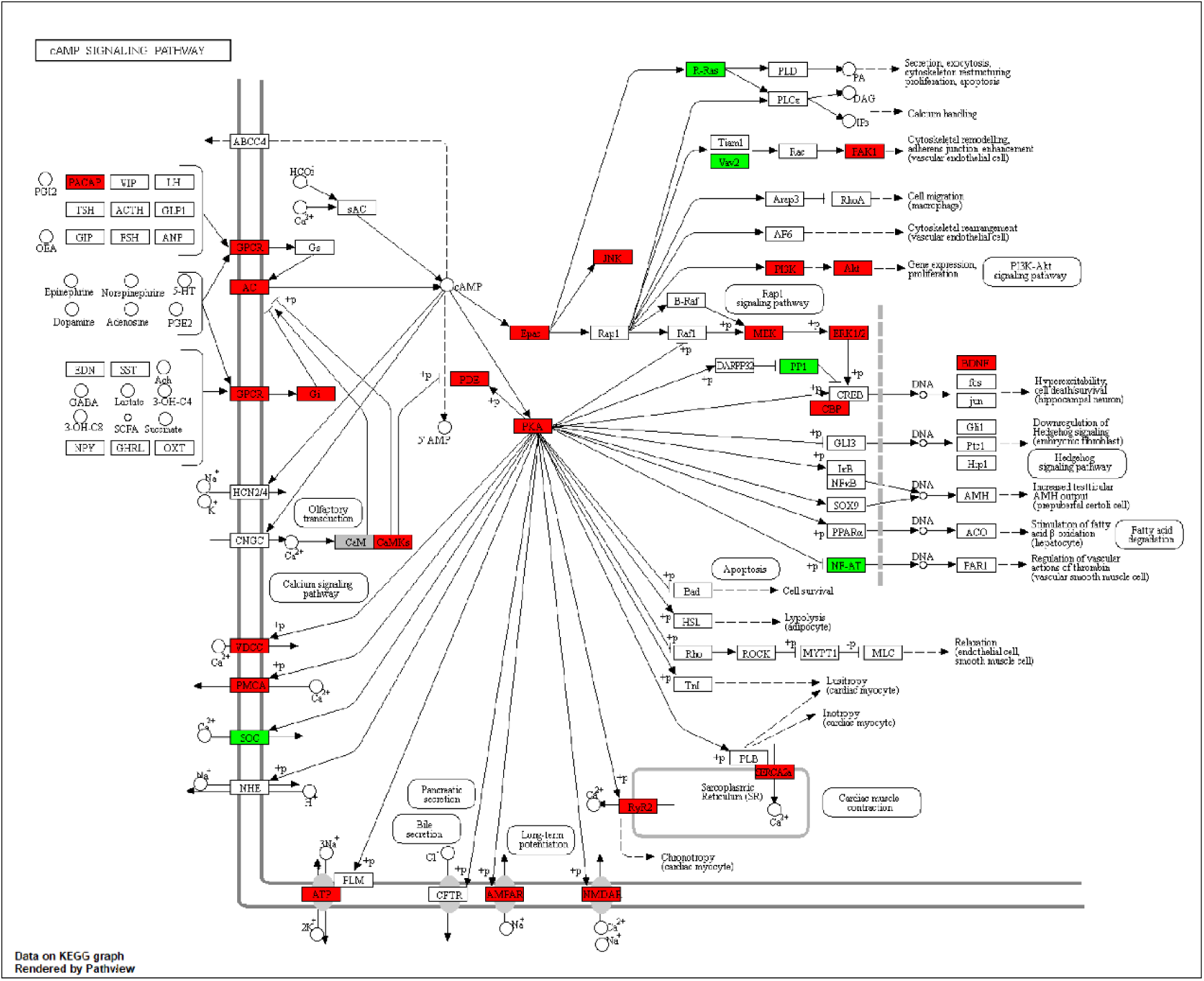
Synapse assembly score correlated genes were involved in cAMP signaling pathway. Graph was downloaded from KEGG (hsa04024). Red: positive correlation genes. Green: negative correlation genes.

**Supplementary Fig. 6.**
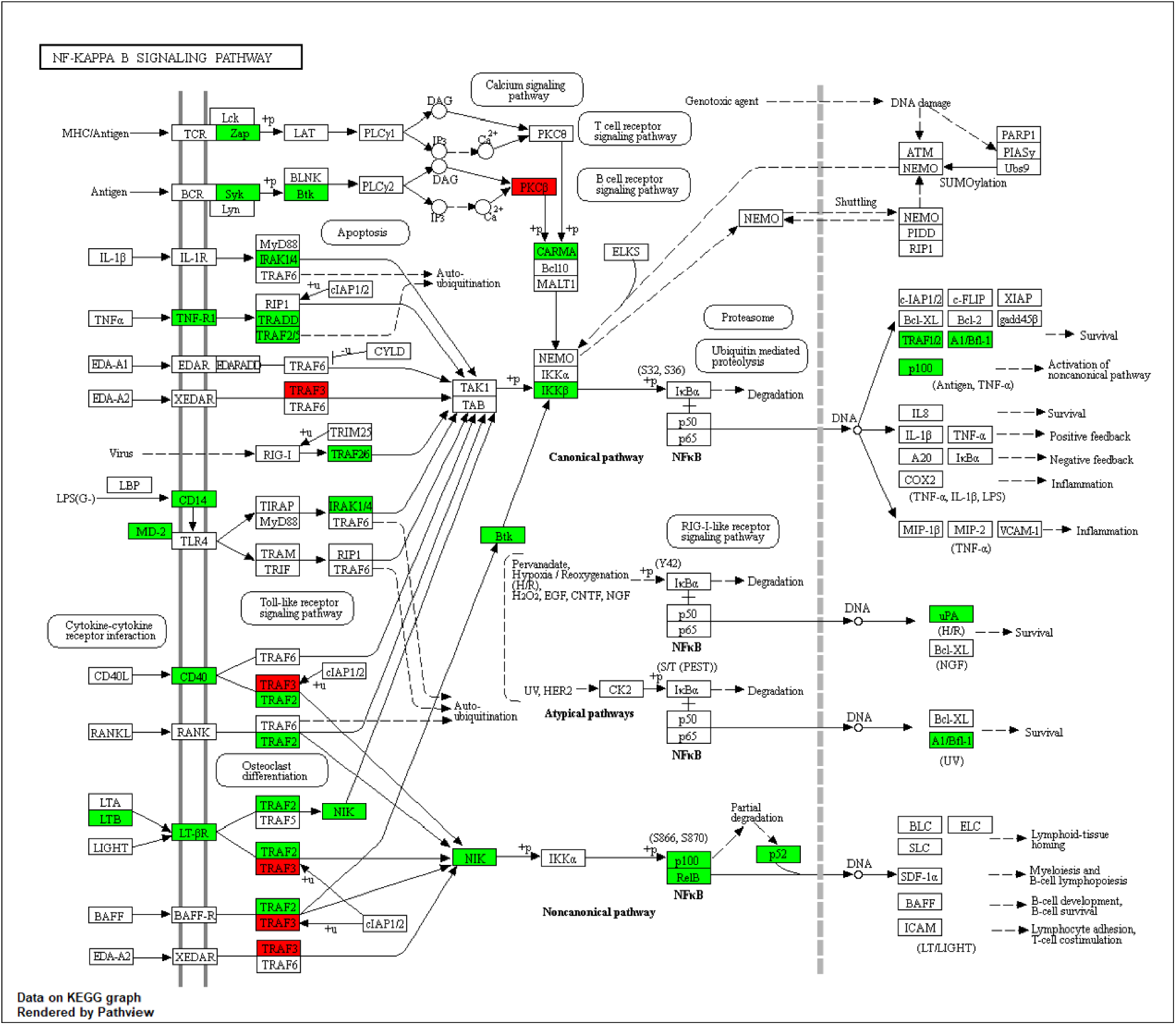
Genes negatively associated with SA score were widely distributed in NF-κB signaling pathway. Graph was obtained from KEGG (hsa04064). Red: positive correlation genes. Green: negative correlation genes.

**Supplementary File 1: List of the 66 SA genes**.

## References

1 Brain and Other Nervous System Cancer - Cancer Stat Facts, <https://seer.cancer.gov/statfacts/html/brain.html> (2019).

2 Ostrom, Q. T. et al. CBTRUS Statistical Report: Primary Brain and Other Central Nervous System Tumors Diagnosed in the United States in 2009-2013. Neuro Oncol 18, v1–v75, doi:10.1093/neuonc/now207 (2016).

3 Louis, D. N. et al. The 2007 WHO classification of tumours of the central nervous system. Acta Neuropathol 114, 97–109, doi:10.1007/s00401-007-0243-4 (2007).

4 Wesseling, P. & Capper, D. WHO 2016 Classification of gliomas. Neuropathol Appl Neurobiol 44, 139–150, doi:10.1111/nan.12432 (2018).

5 Gramatzki, D. et al. Glioblastoma in the Canton of Zurich, Switzerland revisited: 2005 to 2009. Cancer 122, 2206–2215, doi:10.1002/cncr.30023 (2016).

6 Ruda, R. & Soffietti, R. Controversies in management of low-grade gliomas in light of new data from clinical trials. Neuro Oncol 19, 143–144, doi:10.1093/neuonc/now275 (2017).

7 Bolouri, H., Zhao, L. P. & Holland, E. C. Big data visualization identifies the multidimensional molecular landscape of human gliomas. Proc Natl Acad Sci U S A 113, 5394–5399, doi:10.1073/pnas.1601591113 (2016).

8 Cao, H., Wang, F. & Li, X. J. Future Strategies on Glioma Research: From Big Data to the Clinic. Genomics Proteomics Bioinformatics 15, 263–265, doi:10.1016/j.gpb.2017.07.001 (2017).

9 Febbo, P. G. et al. NCCN Task Force report: Evaluating the clinical utility of tumor markers in oncology. J Natl Compr Canc Netw 9 Suppl 5, S1–32; quiz S33, doi:10.6004/jnccn.2011.0137 (2011).

10 Kros, J. M. et al. Circulating glioma biomarkers. Neuro Oncol 17, 343–360, doi:10.1093/neuonc/nou207 (2015).

11 Patel, A. P. et al. Single-cell RNA-seq highlights intratumoral heterogeneity in primary glioblastoma. Science 344, 1396–1401, doi:10.1126/science.1254257 (2014).

12 Tirosh, I. et al. Single-cell RNA-seq supports a developmental hierarchy in human oligodendroglioma. Nature 539, 309–313, doi:10.1038/nature20123 (2016).

13 Venteicher, A. S. et al. Decoupling genetics, lineages, and microenvironment in IDH-mutant gliomas by single-cell RNA-seq. Science 355, doi:10.1126/science.aai8478 (2017).

14 Subramanian, A. et al. Gene set enrichment analysis: a knowledge-based approach for interpreting genome-wide expression profiles. Proc Natl Acad Sci U S A 102, 15545–15550, doi:10.1073/pnas.0506580102 (2005).

15 Barbie, D. A. et al. Systematic RNA interference reveals that oncogenic KRAS-driven cancers require TBK1. Nature 462, 108–112, doi:10.1038/nature08460 (2009).

16 Aibaidula, A. et al. Adult IDH wild-type lower-grade gliomas should be further stratified. Neuro Oncol 19, 1327–1337, doi:10.1093/neuonc/nox078 (2017).

17 Venkataramani, V. et al. Glutamatergic synaptic input to glioma cells drives brain tumour progression. Nature, doi:10.1038/s41586-019-1564-x (2019).

18 Venkatesh, H. S. et al. Electrical and synaptic integration of glioma into neural circuits. Nature, doi:10.1038/s41586-019-1563-y (2019).

19 Zeng, Q. et al. Synaptic proximity enables NMDAR signalling to promote brain metastasis. Nature 573, 526–531, doi:10.1038/s41586-019-1576-6 (2019).

20 Turkalp, Z., Karamchandani, J. & Das, S. IDH mutation in glioma: new insights and promises for the future. JAMA Neurol 71, 1319–1325, doi:10.1001/jamaneurol.2014.1205 (2014).

21 Cancer Genome Atlas Research, N. et al. Comprehensive, Integrative Genomic Analysis of Diffuse Lower-Grade Gliomas. N Engl J Med 372, 2481–2498, doi:10.1056/NEJMoa1402121 (2015).

22 Moslehi, J. J., Salem, J. E., Sosman, J. A., Lebrun-Vignes, B. & Johnson, D. B. Increased reporting of fatal immune checkpoint inhibitor-associated myocarditis. Lancet 391, 933, doi:10.1016/S0140-6736(18)30533-6 (2018).

23 Johnson, D. B. et al. Fulminant Myocarditis with Combination Immune Checkpoint Blockade. N Engl J Med 375, 1749–1755, doi:10.1056/NEJMoa1609214 (2016).

24 Voglis, G. & Tavernarakis, N. The role of synaptic ion channels in synaptic plasticity. EMBO Rep 7, 1104–1110, doi:10.1038/sj.embor.7400830 (2006).

25 Martini, M., De Santis, M. C., Braccini, L., Gulluni, F. & Hirsch, E. PI3K/AKT signaling pathway and cancer: an updated review. Ann Med 46, 372–383, doi:10.3109/07853890.2014.912836 (2014).

26 Snyder, S. H. Neurotransmitters, receptors, and second messengers galore in 40 years. J Neurosci 29, 12717–12721, doi:10.1523/JNEUROSCI.3670-09.2009 (2009).

27 Bozzi, Y. & Borrelli, E. The role of dopamine signaling in epileptogenesis. Front Cell Neurosci 7, 157, doi:10.3389/fncel.2013.00157 (2013).

28 Fimia, G. M. & Sassone-Corsi, P. Cyclic AMP signalling. J Cell Sci 114, 1971–1972 (2001).

29 Sun, S. C. The non-canonical NF-kappaB pathway in immunity and inflammation. Nat Rev Immunol 17, 545–558, doi:10.1038/nri.2017.52 (2017).

30 Zhang, Q., Lenardo, M. J. & Baltimore, D. 30 Years of NF-kappaB: A Blossoming of Relevance to Human Pathobiology. Cell 168, 37–57, doi:10.1016/j.cell.2016.12.012 (2017).

31 Hoesel, B. & Schmid, J. A. The complexity of NF-kappaB signaling in inflammation and cancer. Mol Cancer 12, 86, doi:10.1186/1476-4598-12-86 (2013).

32 Gerlo, S. et al. Cyclic AMP: a selective modulator of NF-kappaB action. Cell Mol Life Sci 68, 3823–3841, doi:10.1007/s00018-011-0757-8 (2011).

33 Berghoff, A. S. et al. Correlation of immune phenotype with IDH mutation in diffuse glioma. Neuro Oncol 19, 1460–1468, doi:10.1093/neuonc/nox054 (2017).

34 Amankulor, N. M. et al. Mutant IDH1 regulates the tumor-associated immune system in gliomas. Genes Dev 31, 774–786, doi:10.1101/gad.294991.116 (2017).

35 Chen, R., Smith-Cohn, M., Cohen, A. L. & Colman, H. Glioma Subclassifications and Their Clinical Significance. Neurotherapeutics 14, 284–297, doi:10.1007/s13311-017-0519-x (2017).

36 Ceccarelli, M. et al. Molecular Profiling Reveals Biologically Discrete Subsets and Pathways of Progression in Diffuse Glioma. Cell 164, 550–563, doi:10.1016/j.cell.2015.12.028 (2016).

37 Yu, G., Wang, L. G., Han, Y. & He, Q. Y. clusterProfiler: an R package for comparing biological themes among gene clusters. OMICS 16, 284–287, doi:10.1089/omi.2011.0118 (2012).

38 Luo, W. & Brouwer, C. Pathview: an R/Bioconductor package for pathway-based data integration and visualization. Bioinformatics 29, 1830–1831, doi:10.1093/bioinformatics/btt285 (2013).

39 Robin, X. et al. pROC: an open-source package for R and S+ to analyze and compare ROC curves. BMC Bioinformatics 12, 77, doi:10.1186/1471-2105-12-77 (2011).

